# Management of Dementia in a Resource-Constrained Sub-Saharan African Setting: Outcome of a Retrospective Survey of Clinical Practice in the Only Neuropsychiatric Facility in Northeastern Nigeria

**DOI:** 10.1101/2024.09.09.24313311

**Authors:** Ibrahim Abdu Wakawa, Umar Baba Musami, Suleiman Hamidu Kwairanga, Placidus Nwankuba Ogualili, Mohammed Yusuf Mahmood, Muhammad Abba Fugu, Mohammed Mala Gimba, Muktar Mohammed Allamin, Zaharudeen Umar Abbas, Muhammad Kawu Sunkani, Zainab Bukar Yaganami, Fatima Mustapha Kadau, Nasir Muhammad Sani, Peter Danmallam, Luka Nanjul, Larema Babazau, Zaid Muhammad, Baba Waru Goni, Babagana Kundi Machina, Celeste M. Karch, Chinedu Udeh-Momoh, Thomas K. Karikari, Chiadi U. Onyike, Mahmoud Bukar Maina

## Abstract

**Introduction:** Dementia prevalence is rising in sub-Saharan Africa due to a combination of factors, including population growth and aging. In resource-constrained settings, such as Northeastern Nigeria, dementia management is challenged by delayed diagnosis and limited specialist care. This study evaluates the burden of dementia and its management at the Federal Neuropsychiatric Hospital Maiduguri (FNHM), the only neuropsychiatric facility in Northeastern Nigeria. The study aims to provide insights into current dementia trends and practices and identify key areas for improvement.

**Methods:** A retrospective analysis of patient records at FNPH Maiduguri was conducted, including patients aged 60 and above diagnosed with dementia between 1999 and 2023. Data on patient demographics, dementia subtypes, comorbidities, symptoms, diagnostic investigations, and treatment modalities were analysed.

**Results:** The Available record from the hospital health records register showed that the total number of diagnosed cases of dementia in the FNHM is 1,216 cases with a male predominance (56%). Alzheimer’s disease was the most common subtype (60.5%), followed by vascular dementia (24.5%). Hypertension was the most frequently reported comorbidity (41.6%). Cognitive symptoms, particularly memory loss, were reported in all cases, while behavioural symptoms, such as agitation and hallucinations, were reported in some cases. The most commonly administered treatments included cognitive enhancers (donepezil), supplements (gingko biloba), and non-drug therapies (psychoeducation). However, 70.9% of patients were lost to follow-up, highlighting a critical gap in long-term care.

**Conclusion:** The increasing burden of dementia at the only neuropsychiatric facility in Northeastern Nigeria highlights the urgent need for investments and targeted interventions. Enhancing patient engagement, strengthening follow-up systems, and expanding diagnostic and treatment capacities will improve care outcomes and address the growing demands for dementia management in this underserved region.

## INTRODUCTION

The global demographic transition carries a concomitant increase in the population of older adults aged 60 years and above, and sub-Saharan Africa is not exempted from this phenomenon. [1–3] From approximately 24 million people aged 60 and older in 1980 to 74 million by 2020, the total number of older Africans is projected to triple between 2020 and 2050. [4] Nigeria, the most populous country on the continent, has the highest number of older adults and the nineteenth highest worldwide. [4] From the neurological and psychogeriatric perspectives, the consequences in terms of increased occurrence of both preclinical and symptomatic dementias are grave.

Depending on the severity, dementia is typically characterized by impairment in multiple domains of cognitive functioning such as affectation of memory, [5] executive dysfunction, impairments in basic and Instrumental Activities of Daily Living (IADL), as well as Behavioural and Psychological Symptoms of Dementia (BPSD). [6–9] In addition to the core features of dementia, psychiatric comorbidities and other medical disorders are also common. [10] Because of the multifaceted nature of ‘dementia’, the impact is felt not only by the patient but also by the primary caregivers and clinicians.[11]

The burden of dementia is disproportionately higher in Low- and Middle-Income Countries (LIMCs), particularly in resource-constrained sub-Saharan Africa.[1] Late presentation and diagnosis, limited access to specialist care, inadequate healthcare infrastructure and resources, and a lack of culturally appropriate interventions represent unique challenges in the management of dementia in these settings.[12,13]

In sub-Saharan Africa, there is generally a lack of information on clinical dementia, largely due to challenges in diagnosis. This is primarily because biomarkers are often unavailable and neuroimaging techniques are rarely used. [1] Even in cases where neuroimaging is accessible, it typically involves only structural imaging, such as magnetic resonance imaging (MRI), while more advanced modalities like Positron Emission Tomography (PET) scanners are scarce and expensive.

The two seminal studies conducted on ‘dementia’ in Nigeria up to date were the Indianapolis-Ibadan Dementia Research Project and the Ibadan Study of Aging, which were conducted over three and two decades ago, respectively. [14–16] Both were community studies that assessed the prevalence and risk factors of dementia among community-dwelling elderly adults. The outcomes of both studies revealed some degrees of variation when compared to findings from Western countries, particularly in Europe and North America. While environmental factors might influence these differences, genetic factors, such as variations in the Apolipoprotein E (APOE) gene and its surroundings, likely play a significant role.[17–19]

In addition to the community studies in Nigeria, numerous hospital-based studies have been conducted, mostly using clinical diagnostic criteria and other validated instruments such as the Mini-Mental State Examination (MMSE) and Montreal Cognitive Assessment (MoCA) to assess the severity of cognitive impairment and the Lawton-Brody Instrument to assess instrumental activities of daily living (IADL).[20–23] The diagnoses of other comorbidities are clinically based depending on the clinicians’ discretion. There is a high tendency that psychiatric comorbidities are likely to be underdiagnosed or not detected completely. Furthermore, the lack of diagnostic biomarkers and neuroimaging techniques limits the validity of the diagnosis made.

This study retrospectively examined the clinical management of dementia in a resource-constrained, sub-Saharan African neuropsychiatric hospital. It seeks to provide insights into the approach taken in such an environment and identify potential areas for improvement in healthcare delivery. The study aimed to analyze the trends in clinical dementia cases among elderly Nigerians (aged 60 and above) who presented at the Federal Neuropsychiatric Hospital Maiduguri from all states of Nigeria since its inception. Additionally, we sought to determine the number of reported cases by local government area, adjusted for population, from Borno State, identify subtypes of dementia diagnosed in the last five years, assess the prevalence of psychiatric comorbidities, review routine laboratory investigations and reported symptoms, and evaluate the medications used for dementia management. Through this, we aimed to enhance the understanding of dementia care in low-resource settings and highlight the unique challenges and opportunities for improving dementia management in our region and in sub-Saharan Africa.

## METHODS AND MATERIALS

### Study Location

This study was conducted at the Psychogeriatric Unit of the Federal Neuropsychiatric Hospital, Maiduguri (FNHM), Nigeria, a specialized institution established in 1999 as the primary neuropsychiatric referral centre for the six states within Nigeria’s Northeast geopolitical zone.

### Study Participants

The study population comprised elderly patients aged 60 years and above who had been managed within the Psychogeriatric Unit of FNHM since its inception in 1999. The study subjects had a diagnosis of dementia by the WHO’s International Classification of Diseases, 10th Revision The ICD-10, F01-F99.

### Study Procedure

This was a retrospective observational analysis of data abstracted from the hospital’s electronic medical records (EMR) system, which provided detailed patient histories, including demographic information, dementia type, MMSE scores, BPSD, comorbidities, and results from baseline laboratory investigations. Paper records (case notes) were manually reviewed to ensure the completeness and accuracy of the collected data and to collect data in instances where EMR data were incomplete or unavailable. However, the rest could not be used due to the inability to trace them, ineligibility (such as missing content), or cases where the diagnosis was not confirmed by a consultant or was not based on ICD criteria. A data abstraction template was designed and built into a data entry application using Kobo toolbox to streamline the data entry process. This tool was formated to contain all the necessary sociodemographic and clinical information. A training program was organized for those involved in data entry in order to ensure uniformity, reliability, and standards. this enabled the study doctors to mine the data from the case notes of the patients and enter them directly into the database. Once the data entry was completed, the dataset was exported as a CSV file for analysis.

### Data Analysis

Descriptive statistics are reported. Data analysis for this study was conducted using Anaconda Navigator and Jupyter Notebook for Python 3.10. Pandas and geopandas were used for data processing and descriptive statistics. Matplotlib and Seaborn’s packages were used to plot the stack histograms, geoplots for the choropleth map, and UpSet plot to explore the intersections of symptoms, comorbidities, diagnostic data and treatments and the Upset data was used to describe correlations.

### Ethical Considerations

The study was conducted in full compliance with ethical standards. Ethical approval was obtained from the National Human Research Ethical Review Committee through the Institution’s Review Board of the FNHM. The study adhered to strict confidentiality protocols, ensuring all patient information was anonymized and securely handled.

## RESULT

### Distribution of Dementia Cases, Trends, and Sociodemographic Characteristics of Patients at FNPH Maiduguri

Since the inception of FNHM, a total of 1,216 dementia cases have been reported in patients aged over 60 years. Among these, 655 (56%) were males with a mean age of 71.4 ± 9.7 years, and 509 (43.7%) were females with a mean age of 72.7 ± 8.3 years. In the last five years, 423 cases were recorded, with a mean age of 71.9 years. Of these recent cases, 234 (55.3%) were males with a mean age of 71.6 years, while 189 (44.7%) were females with a mean age of 72.2 years. As shown in Fig 1A, there has been an upward trend in reported dementia cases over time.

**Figure 1:**
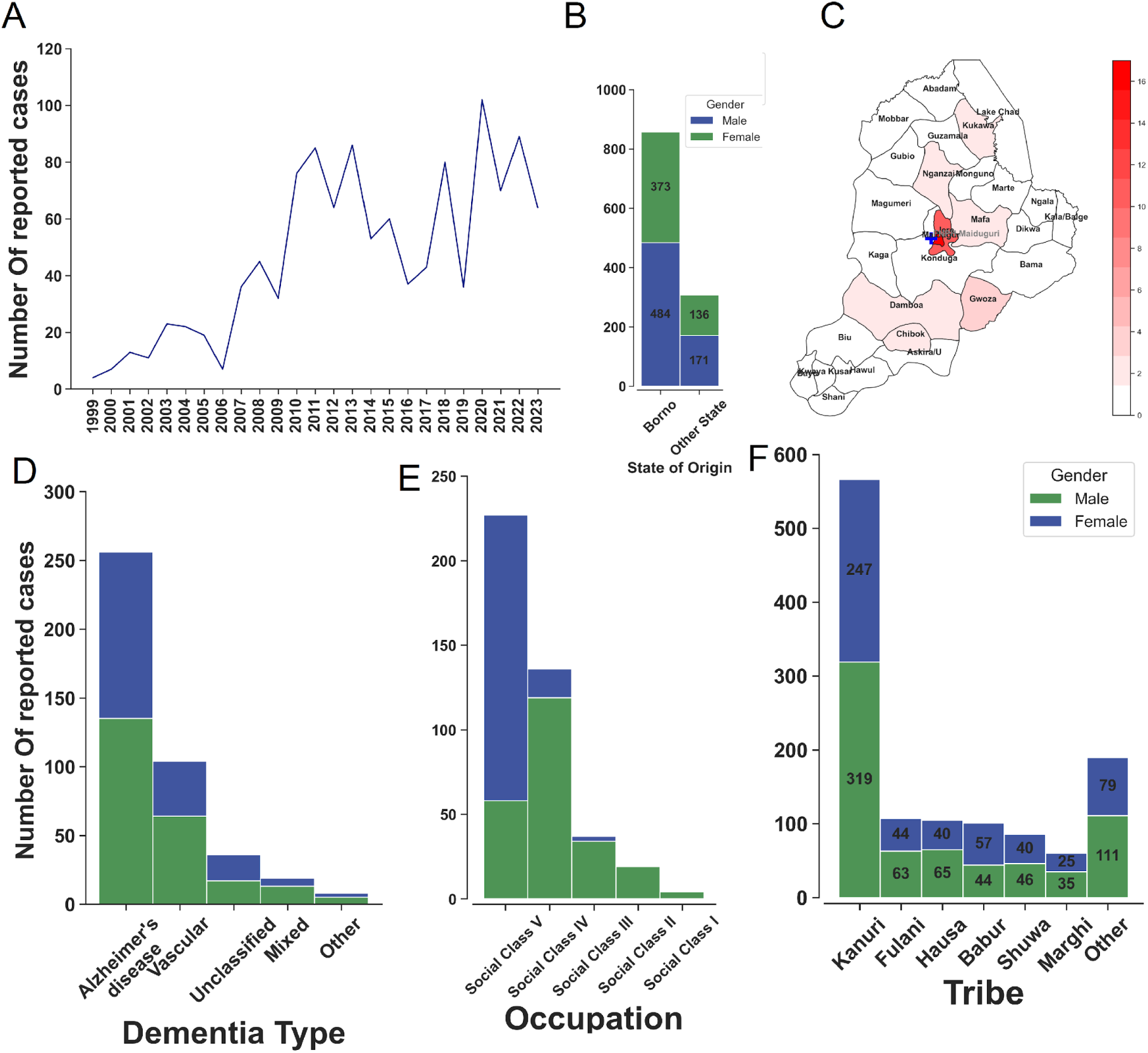
Dementia Case Distribution and Trends at Federal Neuropsychiatry Hospital, Maiduguri (FNHM). (A) The trend of dementia cases at the Psychogeriatric Unit of FNHM from 1999 to 2023. (B) Distribution of patients by state of origin and gender. (C) Choropleth map of Borno State showing the number of reported dementia cases per 100,000 population among patients from Borno State. (D) Distribution of dementia types at the hospital over the last five years. (E) Social class distribution of patients over the last five years. (F) Tribal distribution of patients since the inception of the hospital.

By state of origin, 857 patients (approximately 70%) were from Borno State, making it the most represented state (Fig 1B). Yobe State followed with 130 cases (11.2%), Adamawa State with 49 cases (4.2%), and Bauchi State with 20 cases (1.6%). Jigawa and Sokoto States reported 19 cases (1.6%) and 12 cases (1.0%), respectively. No other state had more than 10 cases. For patients from Borno State, analysis by local government areas (Fig 1C) showed that Maiduguri and Jere LGAs, both urban areas, had the highest number of cases, with 17 cases per 100,000 population in Maiduguri and 10 cases per 100,000 in Jere. No other LGA reported more than two cases per 100,000 population.

Over the last five years, Alzheimer’s disease was the most frequently reported form of dementia, accounting for 60.5% of cases (Fig 1D). Vascular dementia followed 24.5%, and mixed dementia 4.5%. A small proportion of cases were unclassified. Other forms of dementia, including dementia due to head injury, depressive pseudodementia, and frontotemporal dementia, were very uncommon.

The social class distribution of patients over the last five years (Fig 1E) showed that the majority, 53.7%, belonged to Social Class V, consisting of unemployed individuals. This was followed by Social Class IV, with 32.2%, including unskilled workers such as petty traders, subsistence farmers, and messengers. Social Class III, comprising low-skilled workers like junior clerks, drivers, mechanics, and junior military and police personnel, accounted for 8.7% of cases. Social Class II, representing intermediate-skilled professionals like technicians and nurses, has fewer cases (4.5%). Cases were rare in Social Class I, consisting of highly skilled professionals such as doctors, lawyers, and business executives.

Most dementia cases were reported among the Kanuri tribe (Fig 1F), 46.5% of the total. The Fulani tribe followed with 8.8%, the Hausa and Babur with 8.6% and 8.3%, respectively, the Shuwa tribe with 7.1%, and the Marghi with 4.9%. Other tribes collectively accounted for 15.7% of cases.

### Reported Comorbidities and Risk Factors in Dementia Patients Attending FNPH Maiduguri (2019–2023)

Hypertension was the most common comorbidity (Fig 2A), occurring in 41.6% of cases. The frequency of depression was 5.4%, that of stroke was 2.8%, and for diabetes, 0.7%. Parkinson’s disease was observed in 0.9% of cases. Notably, 51.8% of cases had no comorbidity. Correlation analyses showed that 36.2% had hypertension and no other comorbid conditions. Hypertension combined with depression was observed in 2.4% of cases. Among the predisposing factors in dementia patients (Fig 2B), family history was the most prevalent, reported in 8.7% of cases. Other predisposing factors were infrequent. The comorbidities and risk factors among patients with Alzheimer’s disease closely mirrored those seen in the overall dementia population, negating the need for separate analysis and allowing us to focus on dementia management as a whole.

**Figure 2:**
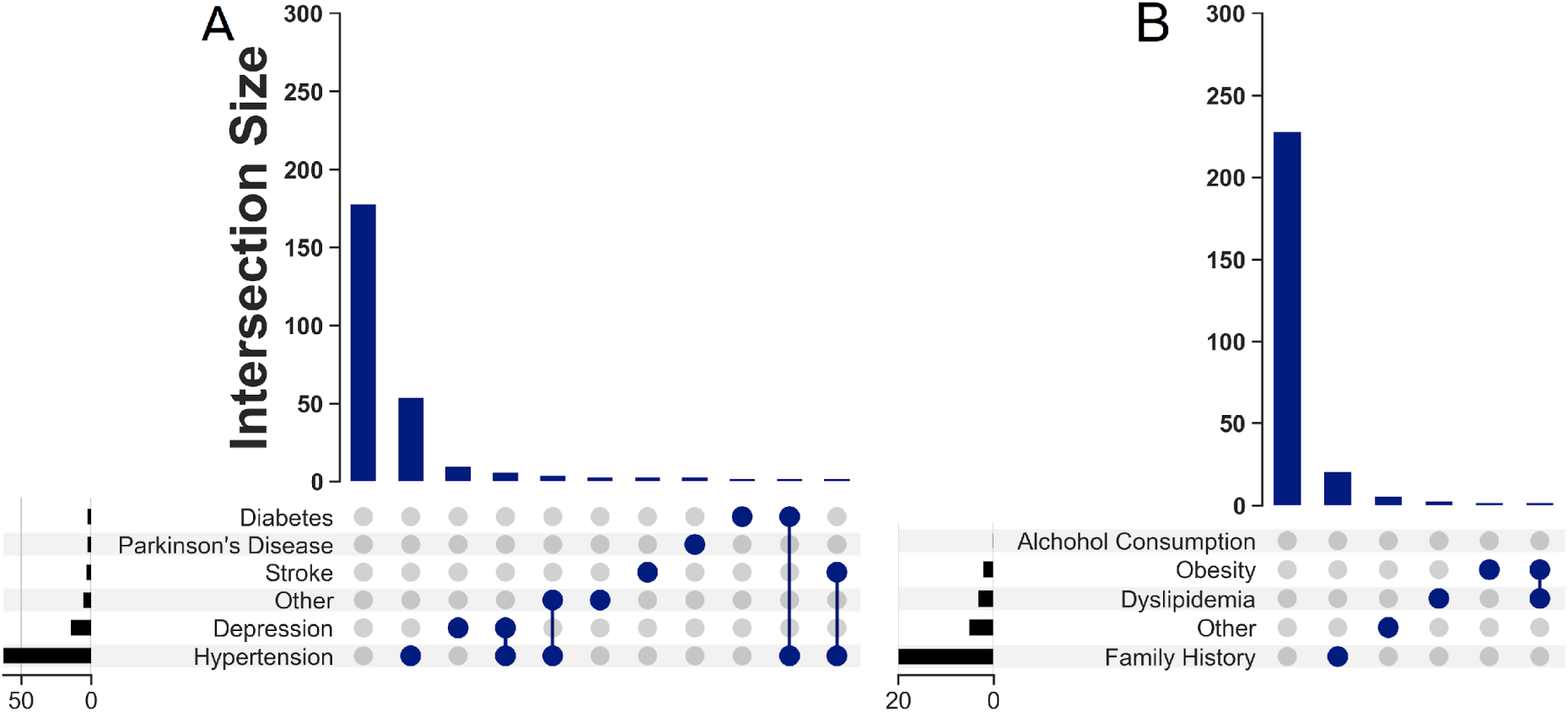
Reported Comorbidities and Risk Factors in Dementia Patients Attending Federal Neuropsychiatric Hospital, Maiduguri (2019–2023). (A) Comorbidities. (B) Predisposing Risk Factors.

### Reported Symptoms and Investigations Conducted in Dementia Patients Attending FNPH Maiduguri (2019–2023)

Among the reported symptoms (Fig 3A), cognitive symptoms were the most prevalent, occurring in all cases. Behavioural symptoms were reported in 77.5%, and neurological symptoms in 11.8%. Cognitive and behavioural symptoms were noted in 66.4% of the cases. A smaller proportion, 21.8%, had only cognitive symptoms. The combination of cognitive, behavioural, and neurological symptoms was observed in 11.1%, while cognitive and neurological symptoms were present in 0.7%.

**Figure 3:**
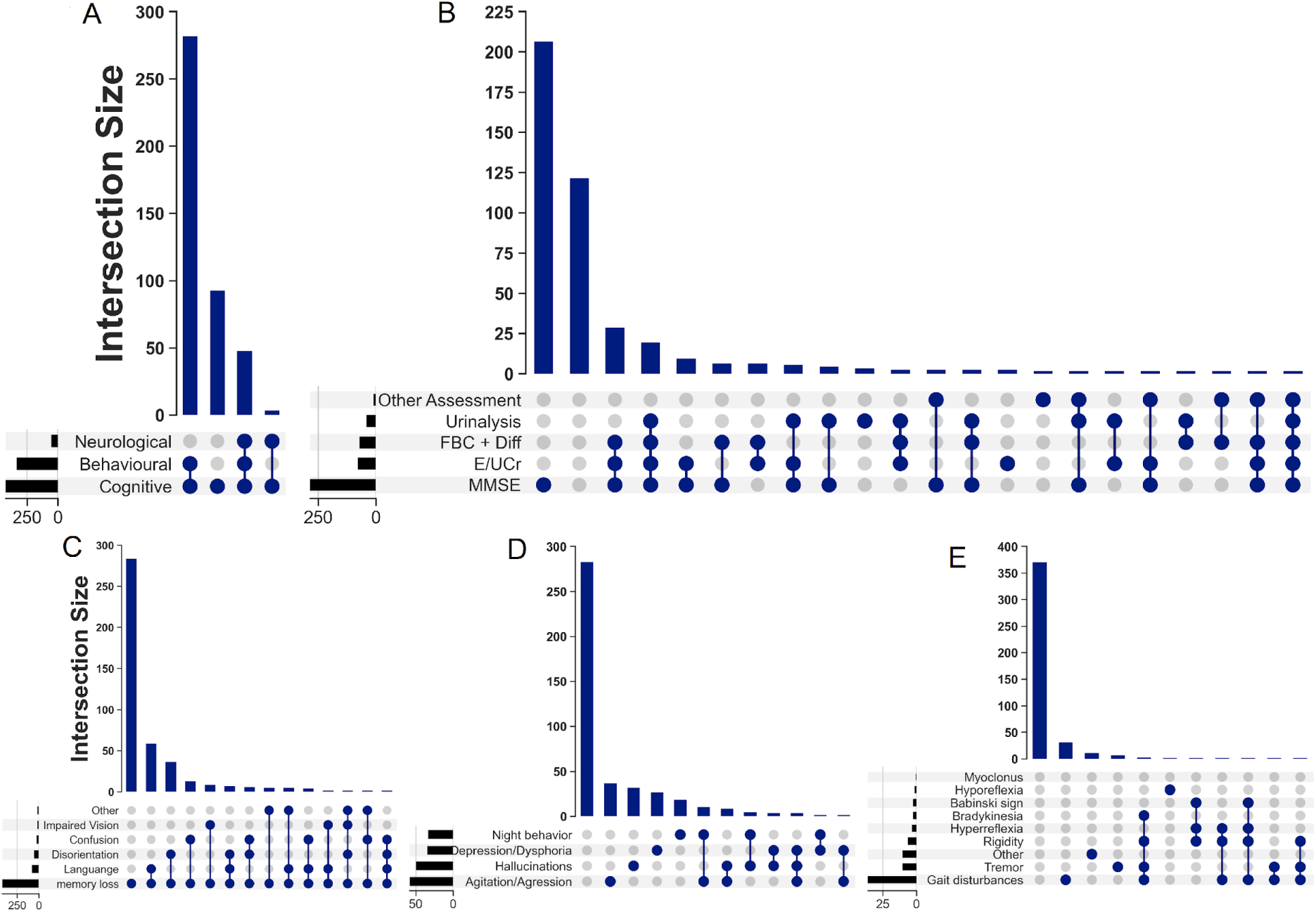
Reported Symptoms and Investigations conducted in dementia patients attending Federal Neuropsychiatric Hospital, Maiduguri (2019-2023). **(A)** Combinations of Reported Symptoms **(B)** Combinations Investigations Conducted **(C)** Combination of Reported Cognitive Symptoms **(D)** Combination of Reported Behavioural Symptoms **(E)** Combination of Reported Neurological Symptoms.

Memory loss was observed in all the cases (Fig 3C). Language difficulties were reported in 17.3%, disorientation in 11.6%, confusion in 5.2%, and impaired vision in 2.4% of cases. In terms of cognitive symptom combinations, 66.9% had memory impairment alone. Memory loss combined with language difficulties was observed in 13.7%, and memory loss combined with disorientation in 8.5%. The combination of memory loss and confusion was noted in 2.8%.

Of the reported behavioural symptoms (Fig 3D), agitation/aggression was the most commonly reported, occurring in 13.7%. Hallucinations were observed in 11.6%, followed by depression/dysphoria in 8.0% and night behaviour disturbances in 7.8%. Notably, the majority, 66.7%, reported no behavioural symptoms. Regarding combinations of behavioural symptoms, agitation/aggression and night behaviour disturbances were reported together in 2.4% of cases.

Of the reported neurological symptoms (Fig 3E), gait disturbances were the most frequently reported, observed in 8.5% of cases. Tremor was observed in 2.4%. Other neurological symptoms were far less common.

The MMSE was performed in 67.4% of cases (Fig 3B). Other routine investigations included electrolytes, urea, and creatinine assays, performed in 17.7%, and Full Blood Count with Differential in 15.8%, and urinalysis in 9.2%. Other tests were less frequently performed.

### Recorded Treatments Given to Dementia Patients Attending FNHM (2019–2023)

Supplements were the most frequently administered of the recorded treatments (Fig 4A), given in 88.4% of cases. Cognitive enhancers were used in 87.2% of cases, while non-drug therapy was employed in 71.4%. Antidepressants were prescribed in 9.7% of cases, and antipsychotics in 5.2%. Regarding treatment combinations, non-drug therapy combined with cognitive enhancers and supplements was administered in 50.1% of cases. The combination of cognitive enhancers and supplements alone was observed in 20.1%. Non-drug therapy and cognitive enhancers together were used in 7.1%, while the combination of cognitive enhancers, supplements, and antidepressants was given in 0.9%.

**Figure 4:**
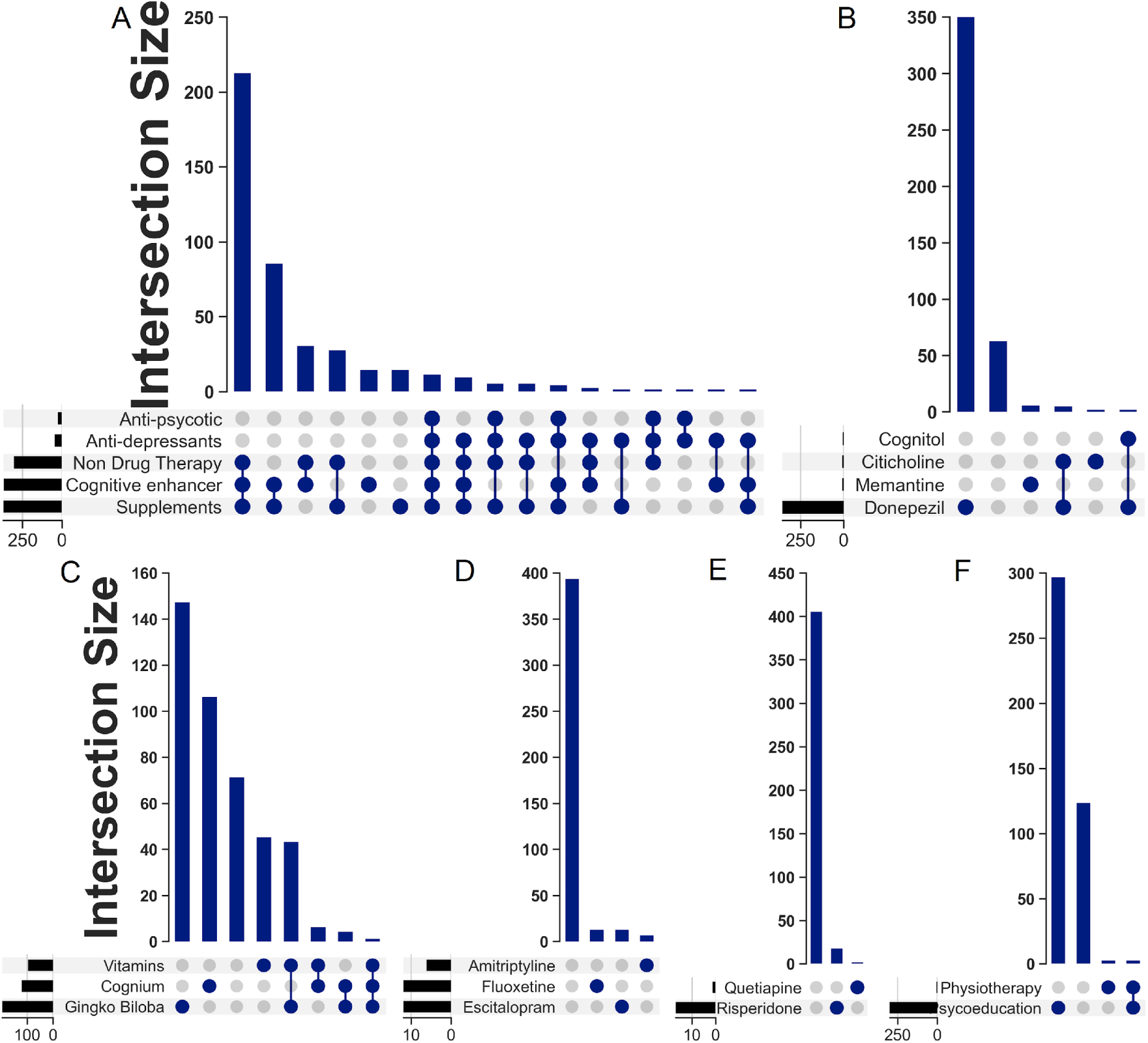
Recorded Treatments Given to Dementia Patients Attending Federal Neuropsychiatric Hospital, Maiduguri (2019–2023). (A) Combinations of Treatment Options, Cognitive Enhancers Administered, (C) Supplements Administered, (D) Antidepressants Administered, and (E) Antipsychotics Administered.

Among the cognitive enhancers, donepezil was the most frequently administered cognitive enhancer (Fig 4B), given in 82.7% of cases. Memantine was rarely prescribed (1.2%), as was citicoline (1 case, 0.2%. The combination of donepezil and citicoline was prescribed infrequently (0.9%), and that of donepezil and vinpocetine in 1 case (0.2%). Notably, 14.7% of cases received no cognitive enhancers. Among the supplements administered (Fig 4C), gingko biloba was the most commonly used in 52.1% of cases. Silk protein hydrolysate (marketed as Cognium) was administered in 31.3% of cases, and vitamins were given in 25.4%. Combinations of supplements were given in 11.5% of cases, while 19.0% did not receive any supplements.

Escitalopram and fluoxetine were the most commonly prescribed antidepressants, each in 29.3%) of cases (Fig 4D). Amitriptyline was prescribed in 14.6% of cases. The remaining patients did not receive any antidepressants. Risperidone was the most commonly administered antipsychotic in 77.3% of cases. Quetiapine was used in just 1 case (4.5%). The remaining patients did not receive any antipsychotic medications.

Of the non-drug therapies (Fig 4F), psychoeducation was the most frequently utilized intervention, reported in 80.1%. Physiotherapy was used in 2 cases (1.7%), while the combination of psychoeducation and physiotherapy was recorded in 2 cases (1.7%). Additionally, 16.5% did not receive any non-drug therapy.

A significant proportion of cases were lost to follow-up, with 70.9% of the cases not returning for further evaluation or treatment. In contrast, 123 patients (29.1%) were confirmed to be alive at the time of the last follow-up.

## DISCUSSION

This study examined the clinical management of dementia at FNHM, focusing on the trends, sociodemographic characteristics, comorbidities, and treatments of patients. Our data showed an upward trend in the frequency of dementia cases, which aligns with global patterns where ageing populations and enhanced diagnostic capabilities have contributed to increased case reporting.[24] This trend suggests that the number of reported cases of dementia is rising due to factors such as increasing life expectancy, prevalent risk factors like hypertension and diabetes, as well as improved awareness and diagnostic methods. The observed dips in case reporting in some years may be linked to external factors, such as the impact of World Health Organization (WHO) interventions such as the Mental Health Gap Action Programme (mhGAP)[25]. During years when the WHO provided free treatment, it is likely that more patients sought care at FNHM, making the facility more accessible compared to other options. In contrast, fewer patients might have reported to the hospital when these interventions were not available, resulting in temporary decreases in case numbers. This pattern highlights the influence of external factors on healthcare utilization and reporting trends, confirming that changes in policy and resource availability can significantly affect the number of reported dementia cases in some regions.

The frequency of dementia among males evaluated at FNM contrasts with the global trend, where females generally exhibit a higher prevalence, particularly of AD. [26]. Similar female dominance has been reported in studies from Uganda, South Africa, and Lagos, Nigeria. [27–29]. However, male-dominant trends have also been observed in other parts of Sub-Saharan Africa, [30,31] potentially due to underreporting among females, cultural factors, and disparities in healthcare access between genders. [31,32].

The significant representation of patients from Borno State may be attributed to the hospital’s status as a major healthcare provider in the region, particularly for dementia cases. This finding aligns with the centralization of specialized healthcare services in urban centres, where facilities like FNHM serve as referral hubs for surrounding areas. [33] The higher prevalence of dementia cases in Maiduguri and Jere Local Government Areas (LGAs), both urban centres, likely reflects better access to healthcare services, leading to earlier and more frequent diagnoses. In contrast, lower rates in rural areas may stem from a combination of limited healthcare access, lower awareness of dementia, and geographic barriers preventing individuals from reaching the hospital.

The predominance of AD among the cases reported at FNHM is consistent with both local and global patterns [26–29,32], where Alzheimer’s is the most common form of dementia. The substantial proportion of vascular dementia cases observed may be linked to the high prevalence of risk factors such as hypertension and diabetes in the region. Similar findings have been reported across Africa, where vascular risk factors are more common, leading to a higher incidence of vascular dementia. [34]

The distribution of dementia cases across social classes at FNHM, with a majority of patients from Social Class V (unemployed), highlights the significant role that socioeconomic factors play in dementia risk. Lower socioeconomic status has been linked to a higher risk of developing dementia[35,36], potentially due to factors such as lower educational attainment, reduced access to healthcare, and greater exposure to environmental risks. These findings are consistent with other studies that emphasize the impact of socioeconomic disparities on dementia incidence. [31,37].

The higher prevalence of dementia among the Kanuri tribe (46.5%) compared to other tribes may be due to their predominant representation in the population of Borno State. This may also reflect cultural and genetic factors that influence dementia prevalence and variations in lifestyle and healthcare access across different tribes. Although the literature on tribal variations in dementia prevalence is limited, the findings at FNHM suggest the need for more research to better understand these differences.

Analyses of the comorbidities and risk factors observed among dementia patients at FNHM from 2019 to 2023 show that hypertension was the most frequently reported comorbidity. This finding aligns with previous literature identifying hypertension as a significant risk factor for dementia, particularly vascular dementia. [38,40] Hypertension contributes to cerebrovascular damage, which can lead to cognitive decline and dementia. [38] It has also been shown to be a risk factor for AD [39]. The high prevalence of hypertension in this study likely reflects broader regional, national and continental trends, where hypertension is a common and often undiagnosed or poorly managed health issue–which exacerbates its impact on cognitive health. [40]

The relatively lower occurrence of other comorbidities, such as depression, stroke, and diabetes, may reflect either underdiagnosis or a genuinely lower prevalence of these conditions in the studied population. Depression, for instance, is a well-known risk factor and comorbidity in dementia, and it can mimic cognitive decline. [41] The low reporting rate could suggest challenges in recognizing and diagnosing depression in dementia patients, a concern highlighted in studies from other resource-constrained settings. [42] Similarly, the small number of cases involving stroke and diabetes—both recognized risk factors for dementia—may point to limitations in screening or reporting practices, which is often an issue in low-resource environments. [43,44]

The fact that over half of the patients had no reported comorbidities raises concerns about the completeness of medical records and/or the thoroughness of patient evaluations. It is possible that some comorbidities were not identified or documented due to limitations in diagnostic capabilities or the prioritization of dementia management over the identification of other health conditions. Another contributing factor could be the overwhelming patient volume at the hospital or across the country, which may restrict doctors’ ability to conduct thorough assessments. In Nigeria, where the doctor-to-patient ratio is critically low, healthcare professionals are often overburdened with large caseloads. [45] There is currently about 1 neurologist per 2 million people in Nigeria. [46] This constraint in the healthcare system can impede the comprehensive management of dementia, as the lack of a full understanding of a patient’s overall health may affect treatment strategies and outcomes. Addressing these systemic challenges is essential for improving the diagnosis and management of dementia and its associated conditions in such high-demand environments.

Regarding predisposing factors, the prominence of family history as a risk factor for AD has long been established in other populations. However, the overall low reporting of other predisposing factors can be attributed to a lack of detailed medical histories or challenges in collecting accurate data. This gap in reporting is consistent with findings from other studies in low-resource settings, where genetic and lifestyle factors are often underexplored due to time constraints, limited patient interaction, or cultural sensitivities surrounding these discussions.[47]

The cognitive profiles described in this study reveal a pattern consistent with both global and local findings. The reporting of memory loss in all cases is consistent with findings globally highlighting memory loss as the most prominent early symptom of dementia. [48–51][52,53] The presence of language difficulties, disorientation, and confusion in a smaller proportion of cases aligns with global observations. For example, language difficulties are often associated with more advanced stages of dementia and are linked to cortical atrophy in regions such as the temporal lobe. [54,55]

The high prevalence of behavioural symptoms is also consistent with the literature on dementia. Behavioural and psychological symptoms of dementia (BPSD), including agitation, aggression, and hallucinations, are well-documented and often challenging to manage. [56–58] Other studies demonstrate that these symptoms not only cause significant distress to patients and caregivers but are also predictors of institutionalisation. [59–61] The fact that a significant portion of patients reported agitation/aggression and hallucinations is typical of the BPSD.

The lower prevalence of neurological symptoms, such as gait disturbances and tremors, reported in our study is in line with other studies where these symptoms are typically less common in the early stages of dementia but may become more pronounced as the disease progresses. Neurological symptoms in dementia are often indicative of more extensive neurodegeneration or advanced disease in conditions like Parkinson’s disease dementia, which is characterized by both cognitive decline and motor symptoms [62].

The combinations of symptoms observed in our cohort, particularly the frequent co-occurrence of cognitive and behavioural symptoms, show the complex nature of dementia. This aligns with the previous studies, which emphasise the fact that patients often present with symptoms that span cognitive, behavioural, and neurological domains, further complicating diagnosis and management.[59,63,64]

The lack of detailed data on investigations conducted at FNHM reflects the resource constraints typical of many sub-Saharan African healthcare settings [34]. The limited access to advanced diagnostic tools like neuroimaging and biomarker assays can lead to a reliance on clinical symptoms for diagnosis, potentially contributing to underdiagnosis or misdiagnosis of dementia subtypes. This might also explain the lower reporting of certain symptoms, which may require more sophisticated clinical approaches to be accurately identified.[34,65]

The frequent administration of cognitive enhancers and supplements at FNHM reflects global trends in dementia care, where these interventions are commonly used to manage symptoms and improve quality of life. [31,32,66] The predominant use of donepezil among cognitive enhancers aligns with its established efficacy in improving cognitive function in AD. [31,67] The limited use of memantine, despite its recognised benefits in moderate to severe AD, is due to resource constraints at FNHM, particularly its cost and limited availability in a low-resource setting. When available, its use may be restricted to cases where the patient’s diagnosis specifically warrants it. Studies show that ginkgo biloba is often utilised for its neuroprotective properties, which can support cognitive function in dementia patients, [68] supporting their extensive use at FNHM.

The conservative use of antidepressants and antipsychotics in this study aligns with the caution recommended in the literature regarding their use in the management of BPSD. Antidepressants such as escitalopram and fluoxetine are prescribed to manage depressive symptoms, which are common in dementia, but their benefits must be carefully weighed against potential adverse effects, particularly in elderly patients. [69] This cautious approach is consistent with findings that highlight the need for careful consideration of the risks and benefits of psychotropic medications in dementia care.

The limited use of antipsychotics, with risperidone being the most commonly administered, reflects best practices that advise minimising the use of these drugs due to their association with increased mortality and cardiovascular adverse events in dementia patients. [69,70] Current guidelines emphasise non-drug approaches as the first line of treatment for BPSD, reserving antipsychotics for severe cases where other interventions have failed. The high prevalence of non-drug therapies, particularly psychoeducation, supports the recommendations in the literature, which highlight the importance of non-pharmacological interventions. Several studies have highlighted the critical role of such therapies in managing BPSD, which are often inadequately addressed by medication alone. [63,71] The reliance on psychoeducation at FNHM reflects an understanding of the value of patient and caregiver education in managing dementia, particularly in settings where access to advanced pharmacological treatments may be limited.

The combination of non-drug therapy, cognitive enhancers, and supplements in more than half of the cases at FNHM aligns with an approach to dementia care that integrates multiple modalities to address the complex needs of patients. For example, some studies discussed the potential synergistic effects of combining different synthetic and natural compound classes to enhance therapeutic efficacy. [72]

We believe that the significant loss to follow-up among dementia patients in this study reflects cultural attitudes, where many Nigerians seek medical care only when symptomatic and often discontinue follow-up once they feel better or if they find alternatives. Additionally, the lack of reporting or documentation of deaths complicates accurate tracking of patient outcomes. These factors, common in LMIC, hinder the ability to assess the effectiveness of treatments and highlight the need for improved patient education, community engagement, and enhanced communication between healthcare providers and families. The resource constraints in Borno state worsened by the Boko haram conflict, have also severely impacted clinical care in the region, [73] including dementia management. Limited healthcare infrastructure, shortage of medical supplies, and reduced access to specialised services lead to delayed diagnosis and poor follow-up, hindering comprehensive dementia care in the region.

## CONCLUSIONS

This study provides important insights into the clinical management of dementia in Northeastern Nigeria, revealing trends in the incidence of dementia cases and the influence of sociodemographic factors such as gender, social class, and ethnicity. The findings highlight the predominant role of AD and vascular dementia in this population, driven by risk factors like hypertension. The use of cognitive enhancers, particularly donepezil, alongside non-drug therapies like psychoeducation, aligns with global dementia care practices, though resource constraints limit the use of other pharmacological treatments such as memantine. The study highlights the challenges for research posed by underdiagnosed comorbidities and significant patient loss to follow-up, which are exacerbated by Nigeria’s low doctor-to-patient ratio and cultural attitudes towards healthcare. Addressing these systemic issues through improved patient education, healthcare infrastructure investments, and better diagnostic tools is essential for enhancing dementia care outcomes in resource-constrained settings.

## RECOMMENDATIONS

1. Implement programmes to educate patients and caregivers on the importance of continuous care and regular follow-up, even in the absence of symptoms. This could include reminders, home visits, or telehealth services to encourage ongoing engagement with healthcare providers.
2. Strengthen communication channels between healthcare facilities and families to ensure that deaths are reported and documented accurately. This may involve training healthcare workers and community leaders to recognise the importance of death certification in the context of public health data collection.
3. Develop community-based interventions that address cultural attitudes towards dementia care, focusing on increasing awareness and reducing stigma. Partnering with local leaders and organisations could help promote a more proactive approach to managing dementia.
4. Allocate more resources to training healthcare providers in dementia care, particularly in the use of non-drug therapies and the management of comorbidities. This could improve the quality of care and patient outcomes in the long term.
5. Encourage further research into the effectiveness of different dementia treatments in resource-constrained settings and invest in better data collection methods to track patient outcomes, treatment efficacy, and mortality rates.

## Data Availability

All data produced in the present study are available upon reasonable request to the authors

## ACKNOWLEDGEMENTS

Funding for this work was provided by the Rainwater Charitable Foundation and the Alzheimer’s Association to Dr. Mahmoud Bukar Maina. We also acknowledge the Northern Nigeria Dementia Research Group, established through this funding, whose contributions supported the completion of this work. Special thanks to the patients, clinicians, and staff at the Federal Neuropsychiatric Hospital Maiduguri for their invaluable contributions to the diagnosis, management, and documentation of dementia cases over the past two decades.

## AUTHOR CONTRIBUTIONS

The study was conceived, organised and supervised by MBM, IAW and CUO. Data curation was done by UBM, SHK, PNO, MYM, MAF, MMG, MMA, ZUA, MKS, ZBY, FMK, NMS, PD and LN managed by MBM and IAW. Data analysis was done by SHK. The paper was written with contributions from all authors. Methodology and study design were guided by BKM, BWG, LB, ZM, CU and TKK. MBM, IAW, CUO, CU and TKK who provided expert guidance and facilitated discussions throughout the project.

